# Trust in the scientific research community predicts intent to comply with COVID-19 prevention measures: An analysis of a large-scale international survey dataset

**DOI:** 10.1101/2021.12.08.21267486

**Authors:** Hyemin Han

## Abstract

In the present study, I explored the relationship between people’s trust in different agents related to prevention of spread of COVID-19 and their compliance with pharmaceutical and non-pharmaceutical preventive measures. The COVIDiSTRESSII Global Survey dataset, which was collected from international samples, was analysed to examine the aforementioned relationship across different countries. For data-driven exploration, network analysis and Bayesian generalized linear model (GLM) analysis were performed. The result from network analysis demonstrated that trust in the scientific research community was most central in the network of trust and compliance. In addition, the outcome from Bayesian GLM analysis indicated that the same factor, trust in the scientific research community, was most fundamental in predicting participants’ intent to comply with both pharmaceutical and non-pharmaceutical preventive measures. I briefly discussed the implications of the findings, the importance of trust in the scientific research community in explaining people’s compliance with measure to prevent spread of COVID-19.

## Introduction

Since the onset of the current COVID-19 pandemic, different agents, including but not limited to, governments, organizations, and scientific communities, have been developing, implementing, and enforcing measures to prevent spread of COVID-19. Such measures embrace both pharmaceutical and non-pharmaceutical means. Since late 2020, there have been several COVID-19 vaccines approved for public use [1]. Even before approval of the first COVID-19 vaccine, diverse non-pharmaceutical measures, such as mask use, social distancing, mandatory self-isolation, stay-at-home order, have been implemented and enforced [2]. Although the pandemic has not concluded, data collected so far suggests that implementation of such preventive measures have significantly contributed to prevention and mitigation of severe COVID-19 outbreaks [3,4].

Given the importance of preventive measures in prevention of spread of COVID-19, whether public is compliant with such measures would be critical in the current pandemic situation [5]. Even if diverse preventive measures that have been found to be effective are planned and implemented by agents, without people’s compliance with the measures, successful control of the pandemic could not be achieved [6]. For instance, rejection of and noncompliance with the recommended and required preventive measures associated with political debates resulted in the recent drastic increase in COVID-19 cases and deaths caused by the Delta variant in multiple countries across the globe [5,7]. Hence, it would be important to understand which factors are involved in people’s compliance as well as noncompliance with preventive measures.

Previous research has suggested that trust in agents addressing pandemic-related matters is one of the most fundamental factors predicting compliance with preventive measures [8]. For instance, several researchers have examined and reported significant association between trust in governmental agents and organizations in the domain of health care (e.g., World Health Organization), and vaccination intent and compliance with non-pharmaceutical preventive measures [9–11]. Furthermore, trust in science and scientific research communities, which play fundamental roles in developing preventive measures and proposing guidelines based on evidence, has also been considered as a central factor in predicting compliance [12,13]. This would be particularly important within the context of the current pandemic, because spread of misinformation and conspiracy theories, which are closely associated with distrust in science and particularly problematic in recent days, drives people’s tendency to disobey mandatory preventive measures and vaccination requirement [14].

Although the aforementioned previous studies have examined the importance of trust in compliance with preventive measures, several limitations would warrant further investigations. First, the majority of the previous studies was conducted with datasets collected from single or a limited number of countries. Given the current COVID-19 pandemic is a global issue [15], it would be necessary to collect data across diverse countries in examining the mechanism of compliance tendency. Such relatively small-scale research based on data from a small number of countries might not be sufficient to draw conclusions that can be well generalizable across the globe.

Second, in terms of methodology, the previous studies employed conventional analysis methods, which are based on frequentist perspective; such conventional methods are suitable to test one specific null hypothesis and/or model, but not ideal for model exploration [16]. For instance, if we are primarily interested which trust factor is central in prediction of compliance tendency, the previous studies employing conventional methods might not be able to address our interest in a complete manner. In fact, exploration of the best prediction model among multiple competing candidate models requires analysis methods specialized in data-driven analysis, in lieu of conventional hypothesis-driven analysis [17]. Thus, the findings from the previous studies that primarily focused on trust in specific agents and used conventional methods would not show us the full picture of how trust in different agents is associated with compliance with different types of preventive measures. Of course, data-driven analysis has limitations, so we need to be careful while employing the approach [41]. Because data-driven analysis is performed without being guided by specific theory, results from the analysis should be interpreted with caution. If a researcher does not refer to relevant theory while interpreting results, the researcher may make a spurious conclusion. Hence, results shall be carefully interpreted while considering their theoretical implications [41]. It would also be desirable to re-test the results from data-driven analysis [42]. For example, a model identified through data-driven analysis might inform additional hypothesis-driven analysis.

## Current Study

In the current study, how people’s trust in different agents predicts their intent to compliance with preventive measures and get vaccinated within the context of the COVID-19 pandemic will be examined in a data-driven manner with a large-scale international survey dataset to address the aforementioned limitations in the prior research. Unlike the previous studies employing conventional analysis methods, which are suitable for one null-hypothesis testing, I intend to explore which trust factor is particularly important in predicting compliance by exploring the large-scale dataset, the COVIDiSTRESSII Global Survey dataset [18], with data-driven analysis methods.

To conduct the data-driven exploration, I plan to implement two novel analysis methods. First, network analysis will be performed to explore how trust in different agents and compliance with different types of preventive measures are associated with each other. In this exploration, I intend to examine which factor is positioned in the most central and influential position in the network [19]. Second, I will explore the best model predicting compliance with different types of preventive measures with Bayesian model exploration [20]. Through this process, all possible candidate regression models in terms of all possible combinations of trust in different agents will be tested, and the most probable model given data will be identified. Finally, based on results from the aforementioned processes employing data-driven methods, I will examine which trust factors are relatively more important in predicting their compliance with preventive measures across different countries. While interpreting the results, I intend to refer to previous studies addressing topics related to trust and compliance to address the previously mentioned limitation of data-driven analysis.

## Methods

### Dataset

In the present study, I analysed the COVIDiSTRESSII Global Survey dataset, which was collected by the COVIDiSTRESS Global Survey Consortium and is available to public via the Open Science Framework (https://osf.io/36tsd). Originally, the data was collected from 20,601 participants from 62 countries. However, as I employed mixed-effects model analysis to include the between-country effect in analysis, to prevent potential convergence issue [8,21], only data collected from countries where 100 or more participants completed the survey was used in the present study. As a result, I analyzed a subset of the data collected from 14,349 participants from 35 countries. Demographics of the participants included in the subset is presented in Table 1.

**Table 1:**
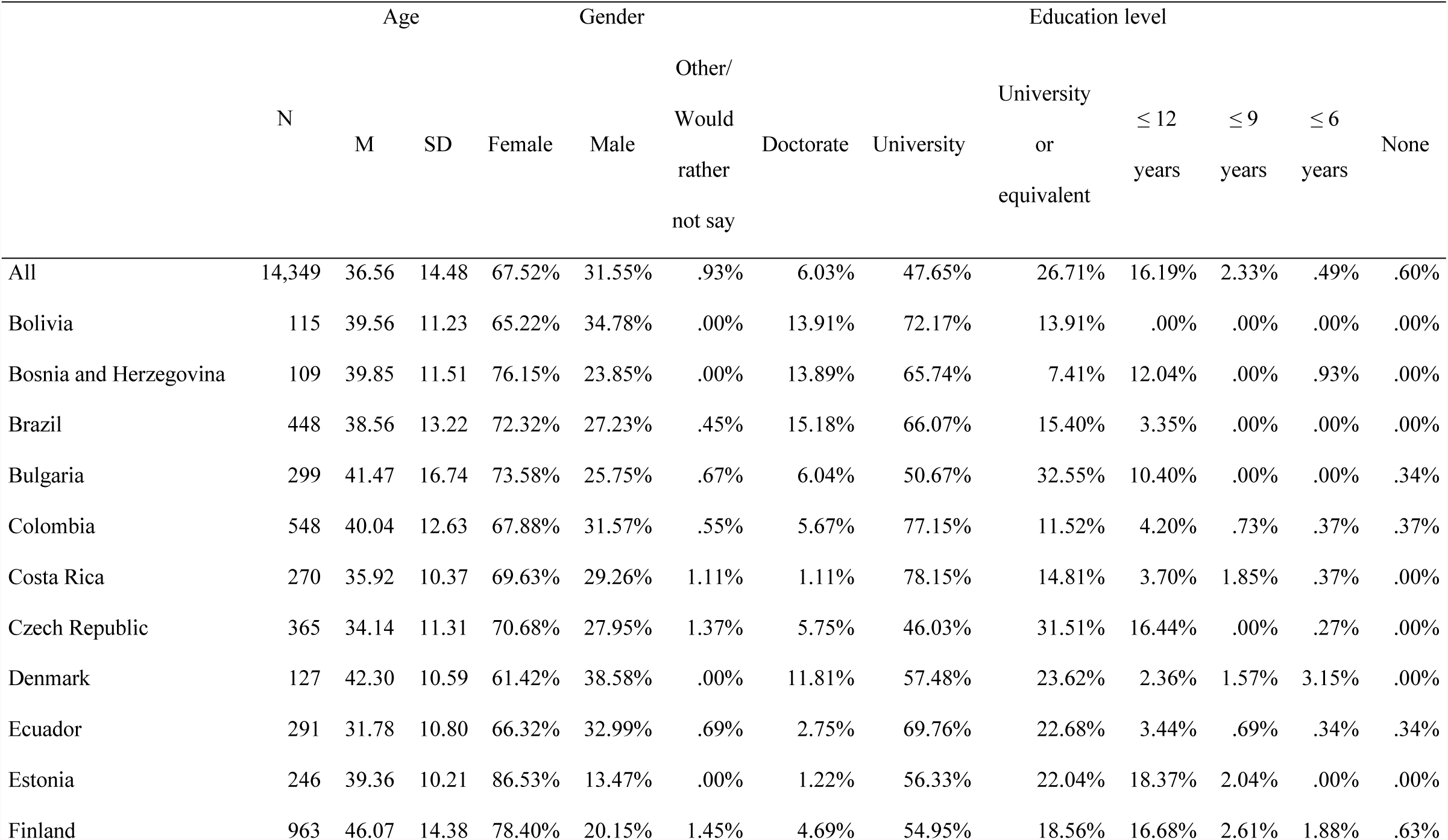
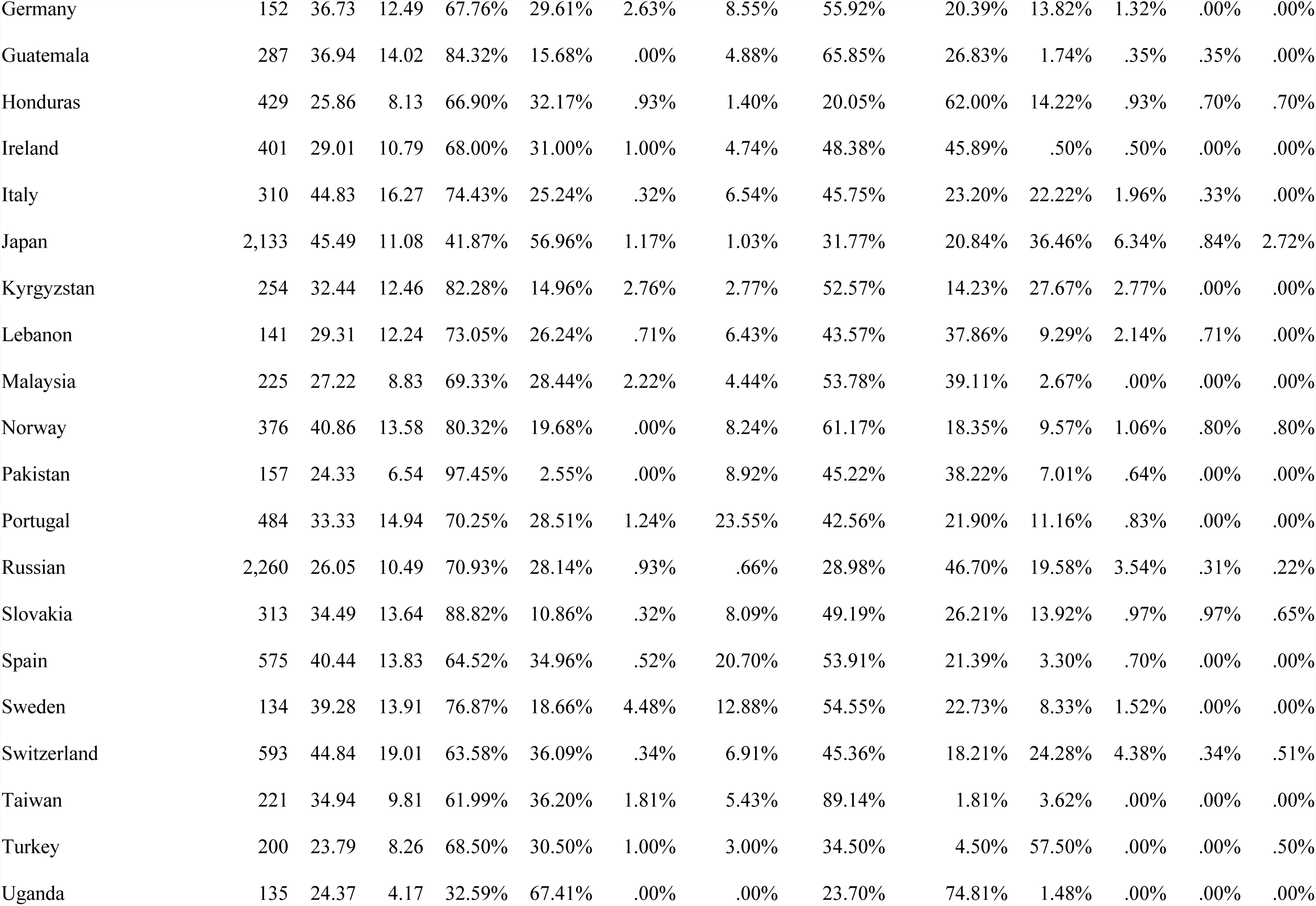
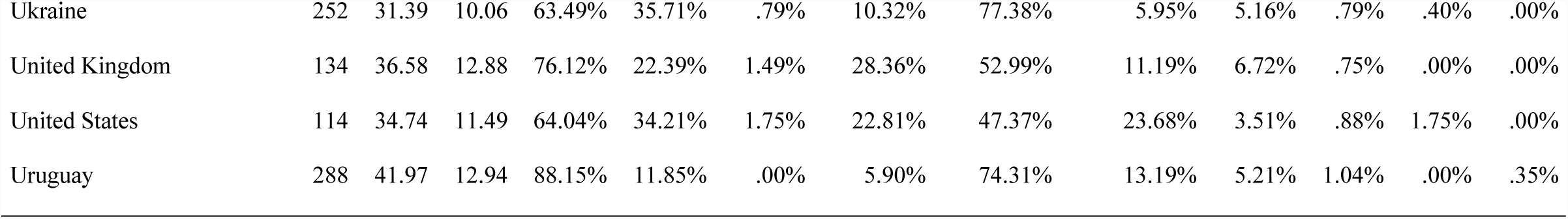
Demographics of the whole dataset and each country.

Further details about data collection and cleaning procedures are explained in the project page (https://osf.io/36tsd). All procedures regarding data collection and informed consent were reviewed and approved by the Research, Enterprise and Engagement Ethical Approval Panel at University of Salford (approval number: 1632) where the project manager of the consortium was affiliated during the data collection period. The author asserts that all procedures contributing to this work comply with the ethical standards of the relevant institutional committee on human experimentation and with the Helsinki Declaration of 1975, as revised in 2008.

### Measures

The employed items were developed by the COVIDiSTRESS Global Survey Consortium members. They were translated and back translated by the consortium members from different countries. Further details about the measures are described in the survey project page (https://osf.io/36tsd).

#### Trust Items

Trust in seven different agents related to development, implementation, and/or enforcement of preventive measures against COVID-19 was surveyed. Participants were asked to what extent they trust each agent based on their general impression. The seven agents were: a parliament or government (Trust 1); police (Trust 2); civic service (Trust 3); health system (Trust 4); WHO (Trust 5); government’s effort to handle Coronavirus (Trust 6); and scientific research community (Trust 7). Participants’ responses were anchored to an eleven-point Likert-type Scale (0: no trust—10: complete trust).

#### Compliance Intent Items

Participants’ intent to comply with eight different types of preventive measures was also surveyed. First, in the domain of pharmaceutical measures, one item, “How willing are you to get the vaccine if one becomes available to you?” was presented to assess their intent to get vaccinated (Compliance 1). Participants’ responses to this item were anchored to five-point Likert scale (1: not willing at all—5: very willing).

Second, in the case of compliance with non-pharmaceutical preventive measures, compliance with seven different types of measures was surveyed. Participants were asked to what extent they were compliant with each measure during the last month. The seven surveyed measures were: washing hands regularly (Compliance 2); wearing a face covering in public when indoors (Compliance 3); wearing a face covering in public when outdoors (Compliance 4); staying at least the recommended distance (Compliance 5); staying at home unless going out for essential reasons (Compliance 6); self-isolating if you suspected that you had been in contact with the virus (Compliance 7); staying away from crowded places generally (Compliance 8). Answers to the items were anchored to a seven-point Likert scale (1: strongly disagree—7: strongly agree).

#### Demographics

Following previous studies examining behavioural and psychological responses to COVID-19 using international survey datasets [8,15,20], several demographic variables were also employed as control variables in the present study. I used participants’ age, gender, and education level in analysis. Participants’ gender was surveyed by presenting three options: female; male; other or would rather not say. The survey presented seven options to ask participants’ education level: PhD or doctorate; university degree (e.g., MA, MSc, BA, BSc); some university or equivalent (still ongoing, or completed a module or more, but did not graduate); up to 12 years of school; up to 9 years of school; up to 6 years of school; none.

### Analysis Plan

#### Network Analysis

To examine the overall association between responses to the seven trust and eight compliance items, I conducted network analysis with *bootnet R* package. The main purpose of network analysis is to demonstrate associations between nodes, trust and compliance in the case of the present study. A connection between two specific node is defined as an edge, which has a weight representing the strength of the association [22]. An edge weight is quantified in term of partial correlation between two nodes by *bootnet*. As an illustrative example, in the case of the edge between Trust 1 and Compliance 1, the edge weight can be understood in terms of correlation between Trust 1 and Compliance 1 after controlling for correlation with all other items in the same network (i.e., Trust 2 … Compliance 8). In a network plot, which visualizes the result of network analysis, an edge between two nodes is presented in the format of a line with a specific thickness, which represents its edge weight, the strength of the association.

While exploring a partial correlation network, *bootnet* employs one technique, graphical LASSO (GLASSO), to identify a regularized partial correlation network through penalizing spurious edge weights [19]. Implementation of GLASSO is required to minimize false positives that may exist in a network of interest. For instance, we can imagine that there is no true non-zero partial correlation between two specific nodes. In the reality, possibly due to noise and/or measurement error, even after controlling for association with other nodes, the edge weight between the two nodes could not exactly become zero, although that is a false positive [19]. Such spurious edge weights can be excluded by GLASSO. Moreover, use of such a penalization method can contribute to prevention of model overfitting [17,23]. Hence, in the present study *bootnet* identified the best network model with the smallest extended Bayesian Information Criterion (EBIC) value to penalize unnecessarily complex and spurious network edge structures.

Once a partial correlation network model was identified with GLASSO, I performed centrality analysis to examine which node located at the most central and influential position in the network. For this purpose, three indicators resulting from centrality analysis, i.e., strength, closeness, and betweenness, were examined for each node [24]. Strength is calculated by summing the absolute values of association strengths, edge weights, of a specific node. Closeness is defined in terms of the inverse of summed distances from one specific node to the other nodes in the same network. Finally, betweenness is estimated in terms of how many times one specific node is in the shortest path between two other nodes in the whole network. In the present study, I examined which node reported the highest strength, closeness, and betweenness values.

#### Bayesian Model Exploration

To examine the best regression model predicting each compliance variable with trust variables, I conducted Bayesian model selection with the Bayesian generalized linear model (GLM) implemented in *BayesFactor* R package. Unlike conventional regression analysis based on frequentist perspective, Bayesian regression analysis enables us to examine to what extent evidence supports a regression model of interest [25]. In the case of conventional regression analysis, only one model can be tested each time, and the resultant *p*-values can only inform us whether a null hypothesis (e.g., whether a null model is the case) shall be rejected [26]. Thus, if our interest is exploration of the best model among all possible candidate models, conventional regression analysis could not be an ideal solution.

Bayesian analysis can provide us with more direct information about whether a specific hypothesis of interest is likely to be accepted given evidence [26]; in the same vein, we can also learn about to what extent a specific model is more likely to be the case compared with other candidate models given evidence as well [27]. Once Bayesian GLM is performed with *BayesFactor*, we can examine the Bayes Factor (BF_M0_) of each model quantifying to what extent the model of interest, Model M, is more strongly supported by data compared with a null model (Model 0) [25]. In the present study, 2log(BF_Mo_) was used for result interpretation. Statistical guidelines suggest that 2log(BF_M0_) ≥ 3 indicates presence of positive evidence supporting Model M against Model 0, 2log(BF_M0_)≥ 6 presence of strong evidence, and 2log(BF_M0_)≥ 10 presence of very strong evidence. When 2log(BF_M0_) < 3, I concluded that evidence is merely trivial or anecdotal [26].

Given the methodological and epistemological benefits of Bayesian analysis, I conducted Bayesian GLM analysis for each compliance dependent variable to identify which trust predictors shall be included in the best regression model [20]. For each dependent variable, I used seven trust variables as candidate predictors, the country as a random effect, and demographic variables as control variables. All trust and compliance variables were standardized at the country level for better convergence, and more straightforward interpretation and comparison of estimated coefficients. Through the process, all possible 128 candidate models, which were created in terms of all possible combinations of seven trust predictors (2^(7+1)^), were estimated and their BF_M0_ were calculated [28]. I identified the best model with the highest BF_M0_ value. Furthermore, I also compared the identified best model and the full model including all seven trust predictors by calculating BF_MF_, a BF value indicating to what extent evidence more strongly supported the best model against the full model. In general, a full model including all candidate predictors is tested and evaluated by resultant *p*-values in conventional regression analysis [16], so I compared the full model with the best model suggested from Bayesian GLM analysis. For both BF_M0_ and BF_MF_, I calculated 2log(BF) values for interpretation. To examine whether the inclusion of the selected covariates significantly altered the outcomes, I performed Bayesian GLM analysis without the covariates. I compared identified best models with results from Bayesian GLM analysis with versus without covariates.

Furthermore, I performed mixed-effects analysis with the indicated best model with *lmerTest* and *brms* R packages. This additional analysis was conducted to examine the effect size of each trust variable included in the best models. Although all predictors included in the best models might be statistically significant in terms of *p*-values, such a significance is perhaps due to a large sample size even if an actual effect is nearly zero or trivial in a practical manner [26]. Effect sizes were calculated in terms of Cohen’s *D* values with *EMAtools* R package after performing multilevel modelling with *lmerTest*. In this process, for each dependent compliance variable, I employed trust variables that were identified to be included in the best models as predictors, the country as a random effect, and demographic variables as control variables. Then, the resultant *D* values were interpreted qualitatively as well as quantitatively. For qualitative interpretation, following [29]’s guidelines, I assumed that a Cohen’s D value within the range of -.10 and +.10 as an indicator of a practically non or trivial effect.

In addition to the qualitative interpretation of effect sizes, I also conducted Bayesian multilevel modelling with *brms* for quantitative interpretation. The same mixed-effects model analysed with *lmerTest* was tested with *brms* for each compliance variable. In this process, I employed the default Cauchy prior, Cauchy (0, 1), suggested by [30] following the previous studies [8,21]. After conducting Bayesian multilevel modelling for each dependent variable, the result was analysed with *bayestestR* package for Bayesian quantitative interpretation of effect sizes. I estimated to what extent the 95% highest density interval (HDI) of the posterior distribution of each trust predictor was within the region of practical equivalence (ROPE) [31,32]. A 95% HDI means an interval that “any parameter value inside the HDI has higher probability density than any value outside the HDI, and the total probability of values in the 95% HDI is 95% [31] (p. 271)”. If 100% of the HDI falls inside the defined ROPE, then the most credible (95%) values of the effect size are completely within the regions of trivial effect, so accepting a null hypothesis (e.g., the effect does not significantly differ from zero) becomes practically reasonable. In this context, a ROPE means a range of a parameter of interest that shall be considered practically near-zero or trivial [32]. Since the guideline that I employed stated that -.10 ≤ *D* ≤ .10 indicates zero or trivial effect [29], I set [-.10 .10] as the ROPE for this quantitative interpretation. By following these steps, I examined to what extent the estimated posterior value of the effect size of each trust predictor in each best model was within the defined ROPE.

## Results

### Network Analysis

Figure 1 is a network plot demonstrating connectivity between seven trust and eight compliance items in the best network model with the lowest EBIC value identified by *bootnet*. Figure 2 shows the result of centrality analysis. In this plot, three centrality indicators, strength, closeness, and betweenness of each node, were presented. All three indicators unequivocally suggest that Trust 7, trust in the scientific research community, is most central in the analysed network.

**Figure 1:**
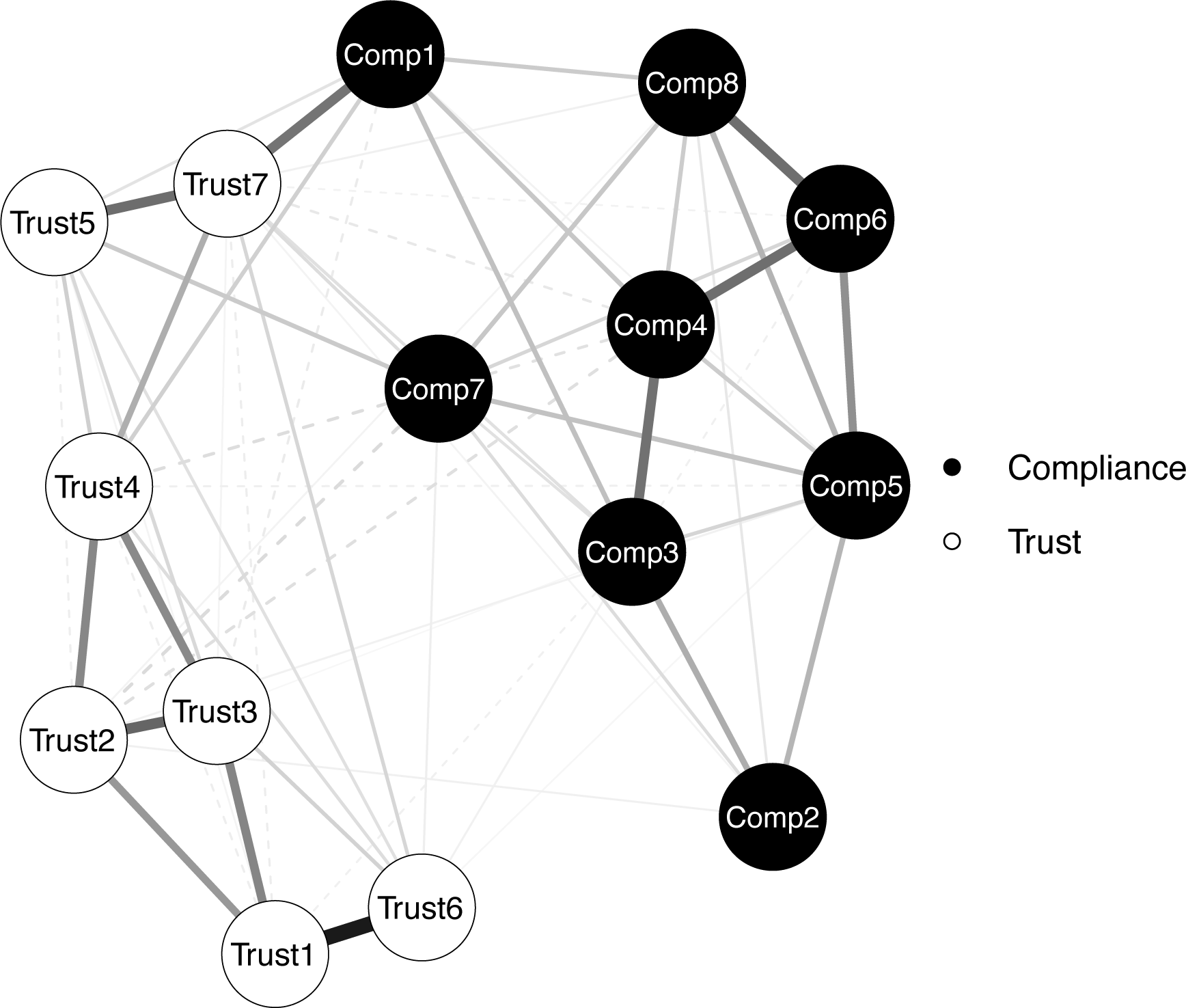
Network plot. Solid line: positive edge weight. Dashed line: negative edge weight.

**Figure 2:**
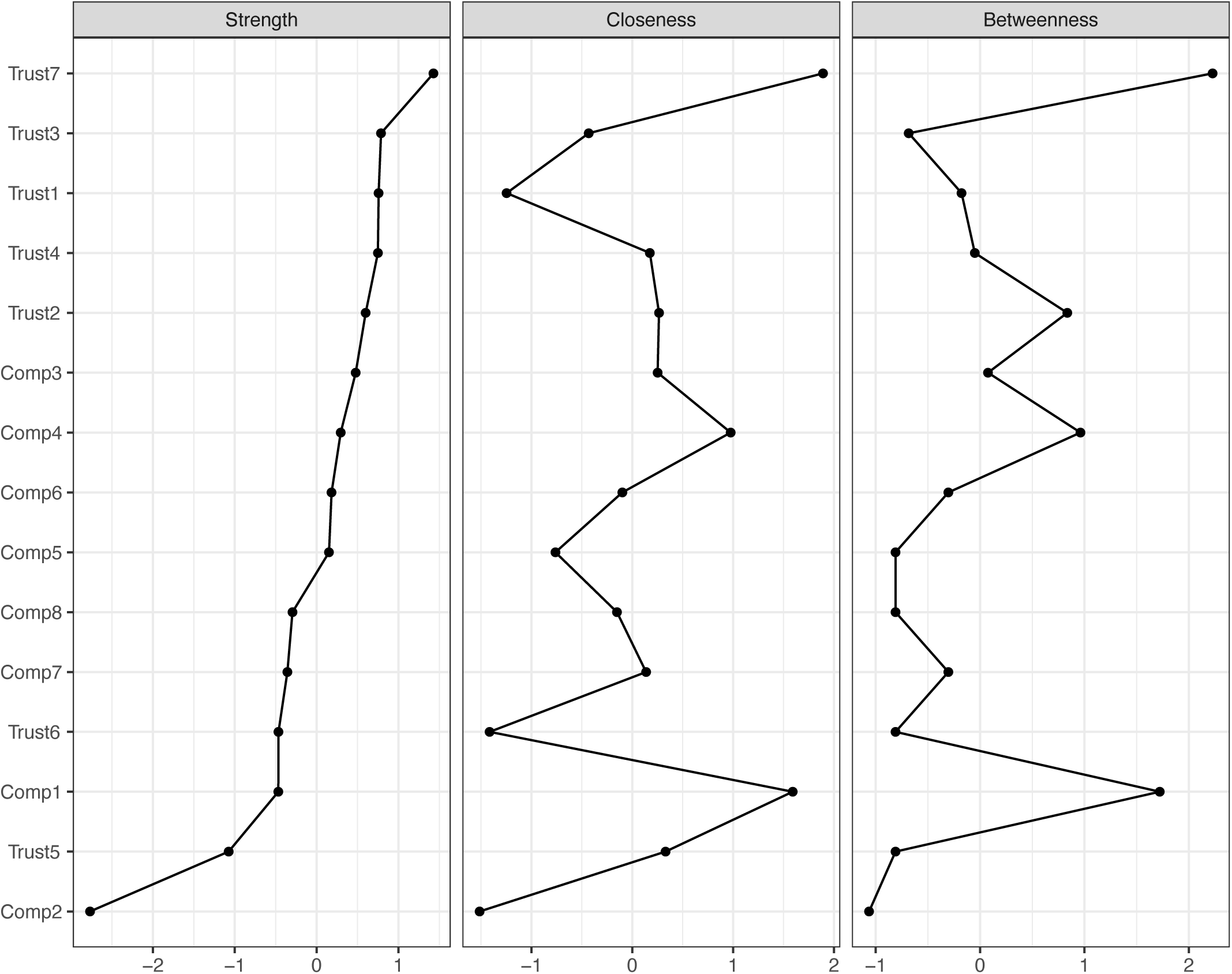
Result from centrality analysis.

### Bayesian Model Exploration

Table 2 demonstrates the results from Bayesian model exploration via Bayesian GLM analysis. The outcome of Bayesian GLM analysis with each compliance dependent variable was presented in each row. Only the coefficients and effect sizes of trust predictors that were included in each best model were presented in Table 2. In addition, the same table reports the proportion of the 95% HDI of each survived trust predictor within the defined ROPE, [-.10 .10].

**Table 2:** Results from Bayesian GLM analysis. Comp. 1—Comp. 8: Compliance 1—Compliance 8. β (SE): Standardized regression coefficient (standard error). ES (ROPE %): Effect size in Cohen’s *D* (% of 95% HDI within the defined region of practical equivalence).

|  | Trust 1 |  | Trust 2 |  | Trust 3 |  | Trust 4 |  | Trust 5 |  | Trust 6 |  | Trust 7 |  | log(Bayes Factor) vs. |  |
| --- | --- | --- | --- | --- | --- | --- | --- | --- | --- | --- | --- | --- | --- | --- | --- | --- |
| | $\beta$ | ES | $\beta$ | ES | $\beta$ | ES | $\beta$ | ES | $\beta$ | ES | $\beta$ | ES | $\beta$ | ES | Null model | Full model |
|  | (SE) | (ROPE %) | (SE) | (ROPE %) | (SE) | (ROPE %) | (SE) | (ROPE %) | (SE) | (ROPE %) | (SE) | (ROPE %) | (SE) | (ROPE %) | (BF<sub>M0</sub>) | (BF<sub>MF</sub>) |
| Comp. 1 | .04 | .06 | -.04 | -.07 | -.04 | -.06 | .08 | .12 | .15 | .25 | .05 | .08 | .27 | .46 |  |  |
|  | (.01) | (100.00%) | (.01) | (100.00%) | (.01) | (100.00%) | (.01) | (99.63%) | (.01) | (.00%) | (.01) | (100.00%) | (.01) | (.00%) | 1375.36 |  |
| Comp. 2 | -.03 | -.04 | .04 | .06 |  |  | .07 | .10 |  |  |  |  | .14 | .25 |  |  |
|  | (.01) | (100.00%) | (.01) | (100.00%) |  |  | (.01) | (100.00%) |  |  |  |  | (.01) | (.00%) | 256.86 | 5.91 |
| Comp. 3 |  |  |  |  |  |  | .03 | .05 | .07 | .11 | .04 | .08 | .17 | .28 |  |  |
|  |  |  |  |  |  |  | (.01) | (100.00%) | (.01) | (100.00%) | (.01) | (100.00%) | (.01) | (.00%) | 480.29 | 5.57 |
| Comp. 4 | .06 | .11 |  |  |  |  |  |  | .07 | .12 |  |  | .11 | .19 |  |  |
|  | (.01) | (100.00%) |  |  |  |  |  |  | (.01) | (100.00%) |  |  | (.01) | (9.10%) | 254.77 | 3.72 |
| Comp. 5 |  |  |  |  |  |  |  |  | .07 | .11 | .05 | .09 | .10 | .17 |  |  |
|  |  |  |  |  |  |  |  |  | (.01) | (100.00%) | (.01) | (100.00%) | (.01) | (30.44%) | 225.27 | 8.82 |
| Comp. 6 |  |  |  |  |  |  |  |  | .06 | .10 | .03 | .05 | .09 | .16 |  |  |
|  |  |  |  |  |  |  |  |  | (.01) | (100.00%) | (.01) | (100.00%) | (.01) | (74.11%) | 158.90 | 9.18 |
| Comp. 7 |  |  |  |  |  |  |  |  | .07 | .12 | .04 | .07 | .11 | .18 |  |  |
|  |  |  |  |  |  |  |  |  | (.01) | (100.00%) | (.01) | (100.00%) | (.01) | (24.57%) | 177.70 | 7.67 |
| Comp. 8 |  |  |  |  |  |  |  |  | .06 | .09 | .03 | .05 | .13 | .22 |  |  |
|  |  |  |  |  |  |  |  |  | (.01) | (100.00%) | (.01) | (100.00%) | (.01) | (.00%) | 227.54 | 10.53 |

In terms of BF_M0_ and BF_MF_, all best models identified by Bayesian GLM analysis, except for the best model predicting vaccination intent (Compliance 1), were supported by very strong evidence compared with both the null and full models. In the case of Compliance 1, the full model including all seven trust predictors was identified as the best model.

When the best models identified with versus without covariates were compared, in the cases of Compliance 1, 3, 4, 6, and 8, there was no significant change. When Compliance 2 was examined, the best model identified without covariates included Trust 2, 4, and 7 as predictors. In the case of Compliance 5, the predictors in the best model without covariates were Trust 2, 5, 6, and 7. When the best model predicting Compliance 7 was explored without covariates, Trust 2, 5, 6, and 7 were identified as predictors. Although the best models changed in these cases, in the cases of Compliance 2 and 7, the originally identified best models with covariates were not significantly worse given 2log(BF) < 3. Only in the case of Compliance 5, the best model identified with covariates was significantly but not very different from that identified without covariates, 2log(BF) = 4.09.

## Discussion

In the present study, first, I conducted network analysis to understand association between participants’ trust in seven different agents addressing the COVID-19 pandemic and their intent to comply with eight different types of preventive measures. Second, Bayesian GLM analysis was performed to explore the best model predicting each compliance intent variable with trust predictors. From network analysis, robust connectivity between trust and compliance variables even after penalizing unnecessary edge features via GLASSO was demonstrated in the visualized network plot. Among all nodes, including both all seven trust and eight compliance variables, Trust 7, trust in the scientific research community, was found to be most central in the network according to all three centrality indicators, strength, closeness, and betweenness. The similar trend was also reported from Bayesian GLM analysis to identify the best model predicting each compliance variable. Although several other trust variables were included in the identified best models, only Trust 7 was included in all eight best prediction models. Such a trend was consistent even when Bayesian GLM analysis was conducted without covariates.

Although I found the significant model change in the case of Compliance 5, the significance of Trust 7 in prediction was consistently supported. Furthermore, in terms of the Cohen’s *D* and proportion of the 95% HDI within the [-.10 .10] ROPE [29,31,32], Trust 7 reported the greatest effect size, which was most likely to be out of the region of near-zero or trivial effect, compared with all six other trust predictors.

The findings suggest that in predicting people’s intent to comply with both pharmaceutical (e.g., vaccination) and non-pharmaceutical measures to prevent spread of COVID-19 (e.g., hand washing, mask use, social distancing, self-isolation), trust in scientific research and the community of scientists play the most fundamental role in the prediction compared with trust in other agents, e.g., government, healthcare system, health organization [12,13]. Given that such measures were primarily tested and suggested by scientific studies with empirical evidence, even if their implementation and enforcement are tasks to be done by other agents, trust in science is expected to make the greatest, fundamental influence on people’s compliance [33]. Hence, if people do not have robust trust in scientific research regarding COVID-19, then they are unlikely to abide by preventive measures implemented by governments and health-related organizations [13].

Given the prevalence of distrust in science, which is being closely linked to widespread of misinformation and conspiracy theories [34], within the current situation, the potential reason of why such distrust contributing to noncompliance with preventive measures would be worth consideration. [35] argued that epistemological doubts and ontological insecurity about scientific knowledge shared within the modern society has promoted and reinforced conspiracy theories and then challenged trust in science among public. A trend related to widespread conspiracy theories and distrust in science is also influential in the current pandemic situation [14]. Conspiracy theories and distrust in science regarding COVID-19 have been promoted by political motive and ideology, authoritarianism, and extremism in particular, and resulted in distrust in scientific evidence supporting preventive measures, and finally, rejection of and noncompliance with recommended preventive measures [34,36]. Furthermore, lack of rational deliberation and reflection upon information and messages is also reported to relate to acceptance of conspiracy theories and distrust in science, which eventually cause noncompliance [37].

Therefore, if researchers and policy makers are interested in promoting people’s compliance with preventive measures, which are suggested and supported by scientific research, they need to consider how to promote people’s trust in scientific research and scientists. Although consideration of concrete solutions for promoting trust in science in public is out of the scope of the current study, let me list a couple of possible starting points. Educators may start with improving science education to educate science-informed citizens who are capable of rationally evaluating and accepting knowledge and information around them in a scientific manner [38]. Moreover, it would be possible to examine how to improve science communication with public, which improve people’s understanding and perception on science [39]. Because many of the current social issues related to noncompliance with COVID-19 preventive measures have been emerged from and reinforced by misinformation shared through diverse forms of media, improvement of science communication would be required to address the issues [40].

We may consider several strengths of the present study and how it could make significant contributions to literature. First, a large-scale international survey dataset, the COVIDiSTRESSII Global Survey dataset, was analysed instead of a relatively small-size dataset collected from a limited number of countries. Because the COVID-19 pandemic is a global issue, findings from the current study will be able to provide researchers and policy makers across the globe with useful insights about how to promote people’s compliance with preventive measures based on generalizable evidence from a cross-national investigation. Second, I explored the overall association between compliance and trust in different agents instead of testing specific hypotheses. With novel quantitative methods, network analysis and Bayesian GLM analysis, I was able to demonstrate that trust in scientific research is most influential and fundamental in predicting compliance.

However, several limitations in the present study may warrant further investigations. First, while collecting data regarding compliance, the project consortium employed the self-report method for the feasibility of the global survey project. Hence, whether the reported compliance intent predicts compliance behaviour in the reality could be questionable. Second, we only employed demographical variables as covariates, although previous research suggested other potential factors, e.g., political orientation, religiosity, significantly associated with people’s trust in science as well as compliance with preventive measures [14]. Third, only one item per trust in each specific agent or compliance with each specific preventive measure was employed in the survey. Because the consortium was not able to use multiple items per construct due to the feasibility issue, the psychometrical aspects of the trust and compliance items could not be tested in a complete manner. Thus, future studies shall employ more direct measures for compliance and additional covariates, and conduct psychometrics tests by employing multiple items per construct to address the limitations in the current study.

## Data Availability

Further details about data collection and cleaning procedures are explained in the project page (https://osf.io/36tsd).

## Data Availability Statement

The data and codes that support the findings of this study are openly available in the Open Science Framework project page (https://doi.org/10.17605/OSF.IO/Y4KGH).

## References

1. Tregoning JS, et al. Progress of the COVID-19 vaccine effort: viruses, vaccines and variants versus efficacy, effectiveness and escape. Nature Reviews Immunology 2021; 21Published online: October 9, 2021.doi:10.1038/s41577-021-00592-1.

2. Perra N. Non-pharmaceutical interventions during the COVID-19 pandemic: A review. Physics Reports 2021; 913Published online: May 2021.doi:10.1016/j.physrep.2021.02.001.

3. Dagan N, et al. BNT162b2 mRNA Covid-19 Vaccine in a Nationwide Mass Vaccination Setting. New England Journal of Medicine 2021; 384Published online: April 15, 2021.doi:10.1056/NEJMoa2101765.

4. Flaxman S, et al. Estimating the effects of non-pharmaceutical interventions on COVID-19 in Europe. Nature 2020; 584Published online: August 13, 2020.doi:10.1038/s41586-020-2405-7.

5. Gao J, Radford BJ. Death by political party: The relationship between COVID-19 deaths and political party affiliation in the United States. World Medical & Health Policy 2021; 13Published online: June 5, 2021.doi:10.1002/wmh3.435.

6. Borchering RK, et al. Modeling of Future COVID-19 Cases, Hospitalizations, and Deaths, by Vaccination Rates and Nonpharmaceutical Intervention Scenarios — United States, April–September 2021. MMWR. Morbidity and Mortality Weekly Report 2021; 70Published online: May 14, 2021.doi:10.15585/mmwr.mm7019e3.

7. Freira L, et al. The interplay between partisanship, forecasted COVID-19 deaths, and support for preventive policies. Humanities and Social Sciences Communications 2021; 8Published online: December 3, 2021.doi:10.1057/s41599-021-00870-2.

8. Lieberoth A, et al. Stress and worry in the 2020 coronavirus pandemic: Relationships to trust and compliance with preventive measures across 48 countries. Royal Society Open Science 2021; 8: 200589.

9. Bargain O, Aminjonov U. Trust and compliance to public health policies in times of COVID-19. Journal of Public Economics 2020; 192Published online: December 2020.doi:10.1016/j.jpubeco.2020.104316.

10. Lalot F, et al. The dangers of distrustful complacency: Low concern and low political trust combine to undermine compliance with governmental restrictions in the emerging Covid-19 pandemic. Group Processes & Intergroup Relations 2020; Published online: October 30, 2020.doi:10.1177/1368430220967986.

11. Chan HF, et al. How confidence in health care systems affects mobility and compliance during the COVID-19 pandemic. PLOS ONE 2020; 15Published online: October 15, 2020.doi:10.1371/journal.pone.0240644.

12. Plohl N, Musil B. Modeling compliance with COVID-19 prevention guidelines: the critical role of trust in science. Psychology, Health & Medicine 2021; 26Published online: January 2, 2021.doi:10.1080/13548506.2020.1772988.

13. Bicchieri C, et al. In science we (should) trust: Expectations and compliance across nine countries during the COVID-19 pandemic. PLOS ONE 2021; 16Published online: June 4, 2021.doi:10.1371/journal.pone.0252892.

14. Rutjens BT, van der Linden S, van der Lee R. Science skepticism in times of COVID-19. Group Processes & Intergroup Relations 2021; 24Published online: February 4, 2021.doi:10.1177/1368430220981415.

15. Yamada Y, et al. COVIDiSTRESS Global Survey dataset on psychological and behavioural consequences of the COVID-19 outbreak. Scientific Data 2021; 8: 3.

16. Han H, Dawson KJ. Improved model exploration for the relationship between moral foundations and moral judgment development using Bayesian Model Averaging. Journal of Moral Education 2021; Published online: 2021.doi:10.1080/03057240.2020.1863774.

17. McNeish DM. Using Lasso for Predictor Selection and to Assuage Overfitting: A Method Long Overlooked in Behavioral Sciences. Multivariate Behavioral Research 2015; 50: 471–484.

18. Blackburn AM, et al. COVIDiSTRESS diverse dataset on psychological and behavioural outcomes one year into the COVID-19 pandemic. 2021.doi: 10.31219/osf.io/428pz

19. Epskamp S, Fried EI. A tutorial on regularized partial correlation networks. Psychological Methods 2018; 23Published online: December 2018.doi:10.1037/met0000167.

20. Han H. Exploring the association between compliance with measures to prevent the spread of COVID-19 and big five traits with Bayesian generalized linear model. Personality and Individual Differences 2021; 176: 110787.

21. Rachev NR, et al. Replicating the Disease framing problem during the 2020 COVID-19 pandemic: A study of stress, worry, trust, and choice under risk. PLOS ONE 2021; 16Published online: September 10, 2021.doi:10.1371/journal.pone.0257151.

22. Dalege J, et al. Network Analysis on Attitudes. Social Psychological and Personality Science 2017; 8Published online: July 10, 2017.doi:10.1177/1948550617709827.

23. Han H, Dawson KJ. Applying elastic-net regression to identify the best models predicting changes in civic purpose during the emerging adulthood. Journal of Adolescence 2021; 93Published online: December 2021.doi:10.1016/j.adolescence.2021.09.011.

24. Opsahl T, Agneessens F, Skvoretz J. Node centrality in weighted networks: Generalizing degree and shortest paths. Social Networks 2010; 32Published online: July 2010.doi:10.1016/j.socnet.2010.03.006.

25. Wagenmakers E-J, et al. Bayesian inference for psychology. Part II: Example applications with JASP. Psychonomic Bulletin & Review 2018; 25: 58–76.

26. Han H, Park J, Thoma SJ. Why do we need to employ Bayesian statistics and how can we employ it in studies of moral education?: With practical guidelines to use JASP for educators and researchers. Journal of Moral Education 2018; 47: 519–537.

27. Dawson KJ, Han H, Choi YR. How are moral foundations associated with empathic traits and moral identity? Current Psychology 2021; Published online: October 12, 2021.doi:10.1007/s12144-021-02372-5.

28. Gronau QF, et al. A Simple Method for Comparing Complex Models: Bayesian Model Comparison for Hierarchical Multinomial Processing Tree Models using Warp-III Bridge Sampling. 2017.

29. Funder DC, Ozer DJ. Evaluating Effect Size in Psychological Research: Sense and Nonsense. Advances in Methods and Practices in Psychological Science 2019; 2Published online: June 8, 2019.doi:10.1177/2515245919847202.

30. Rouder JN, Morey RD. Default Bayes Factors for Model Selection in Regression. Multivariate Behavioral Research 2012; 47: 877–903.

31. Kruschke JK. Rejecting or Accepting Parameter Values in Bayesian Estimation. Advances in Methods and Practices in Psychological Science 2018; 1Published online: June 8, 2018.doi:10.1177/2515245918771304.

32. Makowski D, Ben-Shachar M, Lüdecke D. bayestestR: Describing Effects and their Uncertainty, Existence and Significance within the Bayesian Framework. Journal of Open Source Software 2019; 4Published online: August 13, 2019.doi:10.21105/joss.01541.

33. Nicola M, et al. Evidence based management guideline for the COVID-19 pandemic -Review article. International Journal of Surgery 2020; 77Published online: May 2020.doi:10.1016/j.ijsu.2020.04.001.

34. Miloševic Đorᄜevic J, et al. Links between conspiracy beliefs, vaccine knowledge, and trust: Anti-vaccine behavior of Serbian adults. Social Science & Medicine 2021; 277Published online: May 2021.doi:10.1016/j.socscimed.2021.113930.

35. Aupers S. ‘Trust no one’: Modernization, paranoia and conspiracy culture. European Journal of Communication 2012; 27Published online: March 29, 2012.doi:10.1177/0267323111433566.

36. Tonkovic M, et al. Who Believes in COVID-19 Conspiracy Theories in Croatia? Prevalence and Predictors of Conspiracy Beliefs. Frontiers in Psychology 2021; 12Published online: June 18, 2021.doi:10.3389/fpsyg.2021.643568.

37. Sutton RM, Douglas KM. Conspiracy theories and the conspiracy mindset: implications for political ideology. Current Opinion in Behavioral Sciences 2020; 34Published online: August 2020.doi:10.1016/j.cobeha.2020.02.015.

38. Fensham PJ. Scepticism and trust: two counterpoint essentials in science education for complex socio-scientific issues. Cultural Studies of Science Education 2014; 9Published online: September 6, 2014.doi:10.1007/s11422-013-9560-1.

39. Bubela T, et al. Science communication reconsidered. Nature Biotechnology 2009; Published online: June 2009.doi:10.1038/nbt0609-514.

40. Mheidly N, Fares J. Leveraging media and health communication strategies to overcome the COVID-19 infodemic. Journal of Public Health Policy 2020; 41Published online: December 21, 2020.doi:10.1057/s41271-020-00247-w.

41. Mazzocchi F. Could Big Data be the end of theory in science? A few remarks on the epistemology of data-driven science. EMBO Reports 2015; 16: 1250–1255.

42. Raghu VK, et al. Integrated Theory- and Data-driven Feature Selection in Gene Expression Data Analysis. 2017 IEEE 33rd International Conference on Data Engineering (ICDE) 2017.doi: 10.1109/ICDE.2017.223

